# USPET: UNSUPERVISED SEGMENTATION OF PET IMAGES

**DOI:** 10.64898/2025.12.15.25342254

**Authors:** Maria K. Jaakkola, Henri Kärpijoki, Teemu Saari, Oona Rainio, Anting Li, Juhani Knuuti, Kirsi A. Virtanen, Riku Klén

## Abstract

**Background:** Segmentation is a routine, yet time-consuming and subjective step in the analysis of positron emission tomography (PET) images. Automatic methods to do it have been suggested, but recent method development has focused on supervised approaches. The previously published unsupervised segmentation methods for PET images are outdated for the arising dynamic human total-body PET images now enabled by the evolving scanner technology.

**Methods:** In this study, we introduce an unsupervised general purpose automatic segmentation method for modern PET images consisting of tens of millions of voxels. We provide its implementation in an easy-to-use format and demonstrate its performance on two datasets of real human total-body images scanned using different radiotracers.

**Results and conclusions:** Our results show that the suggested method can identify functionally distinct areas within the anatomical organs. Combined with anatomical segments obtained from other imaging modalities, this enables great potential to improve clinically meaningful segmentation and reduce time-consuming manual work.

## 1. Introduction

Segmentation is a routine step in the analysis and inspection of positron emission tomography (PET) images. Typically, volumes of interest (VOIs) are drawn manually using visual assessment of PET images combined with structural information provided by additional image modality such as magnetic resonance imaging (MRI) or computed tomography (CT). However, this approach often overlooks areas that are anatomically similar in MRI or CT, but functionally distinct in PET. In addition, manual segmentation is time-consuming and subjective. Thus, several automatic segmentation tools for PET images have been developed.

Recent method development related to automatic segmentation of PET images has heavily focused on supervised deep learning methods based on training data [1, 2]. Notably, here we consider training data based methods as supervised machine learning approaches, despite them not needing any supervision from the end-user. While such tools can be very powerful, their use is limited by the utilised training data, and they often serve a specialised purpose. Common examples are identifying predefined organs [3, 4, 5] or finding tumours [6, 7] from PET images. Yousefirizi *et al*. have provided a clear overview of the strengths and weaknesses of supervised segmentation methods compared to unsupervised ones [8], highlighting the need for both types of segmentation methods. The development of unsupervised segmentation methods has been less active. In 2020 Cui *et al*. published a sophisticated pipeline that includes supervised and unsupervised steps to identify tumour lesions from total body PET images scanned using [^18^F]fluorodeoxyglucose ([^18^F]-FDG) [9]. In addition, in 2019 Vogel *et al*. introduced an unsupervised data-driven approach to identify different brain regions from 3D brain [^18^F]AV145 PET images [10]. Also, the work by Xu *et al*. published in 2018 combines segmentation with different preprocessing steps [11]. Many older approaches have also been introduced in the literature [12, 13, 14, 15, 16]. Although unsupervised methods are not restricted by their training data, the recent methods mentioned here still focus on limited areas like a brain or specialised segments like tumor lesions.

There are several challenges related to the mostly very old methodology for unsupervised segmentation. Two of the most critical problems are 1) the lack of available implementations and 2) the unsuitability for modern images provided by the new total-body PET scanners. In addition to preventing clinical application, the lack of available implementations also hinders further method development, as comparing new tools to old ones is not feasible. The lack of available implementations of the introduced finalised tools applies to both supervised and unsupervised methods. The rise of PET scanners with very large fieldof-view that analyse almost the entire human body at once enables studying interactions between different organs, but on the down side, the output images are so large that many previously used segmentation approaches become computationally infeasible [17].

Here we provide an unsupervised general-purpose segmentation method that can analyse modern large field-of-view PET images. The segmentation pipeline is called USPET for Unsupervised Segmentation of PET images and its implementation is freely available at https://github.com/rklen/USPET. We demonstrate USPET’s performance using two real datasets of human total-body PET images scanned using radiotracers [^18^F]-FDG and [^15^O]H_2_O. Notably, we did not find a ready-to-use publicly available implementations of any supervised or unsupervised tool similar to USPET for comparison.

## 2. Materials and methods

### 2.1. Data

In this study, we utilised two datasets of dynamic totalbody human PET images scanned using different tracers. All participants gave their written consent.

The first dataset is called FDG data in this study. The dataset included dynamic PET images with [^18^F]-FDG as a tracer with a mean dose of 107 MBq (standard deviation 4 MBq). The dataset consisted of 22 PET scans of healthy subjects. The scans had 13 time frames (1·60 s, 6·30 s, 1·60 s, 3·300 s, 2·600 s). We manually segmented the kidneys, lungs, heart, aorta, liver, and brain from these images, see Section 2.3 for further details. The reference number for the ethical committee decision related to this dataset is 14/1801/2022 (The wellbeing services county of Southwest Finland).

The second dataset contained 77 total-body dynamic [^15^O]H_2_O PET scans of patients with suspected coronary artery disease. The scans consisted of 24 time frames (14 · 5 s, 3 · 10 s, 3 · 20 s, 4 · 30 s) and the mean dose injected was 352 MBq with a standard deviation of 24 MBq. The scans used in our analyses were performed during adenosine stress. The reference number for the relevant ethical committee decision was 22/1801/2022. Here this dataset is further referred to as H_2_O data.

### 2.2. USPET

The default preprocessing steps in the USPET pipeline are background detection, denoising, log transformation, scaling, and dimensionality reduction, but the end user can disable all or some of them. In addition, the end user can define how much weight to give to the location of each voxel compared to its intensity over time. We have previously evaluated different preprocessing approaches, and according to those observations, USPET utilises gaussian filtering for denoising, z-score normalisation, and principal component analysis for dimensionality reduction. The built-in background detection initially labels all voxels with mean intensity over time points greater than the average as foreground. Then small foreground bubbles in the background are removed, and the same is done for small background areas within the foreground. Log transformation is done prior to normalisation to avoid bias caused by some peak values. After preprocessing, the actual segmentation is done by first clustering the preprocessed foreground voxels with either k-means or Gaussian mixture model (GMM), and then performing connected component analysis for the clusters to further utilise location information in the segmentation. However, this typically generates too many tiny segments. In postprocessing the small segments are melted into their surrounding segment and the remaining segment labels (integers) are re-organised so that the segment with highest mean intensity in the original image will get the highest label. The background is always labelled with 0. Figure 1 visualises the USPET pipeline.

**Figure 1.**
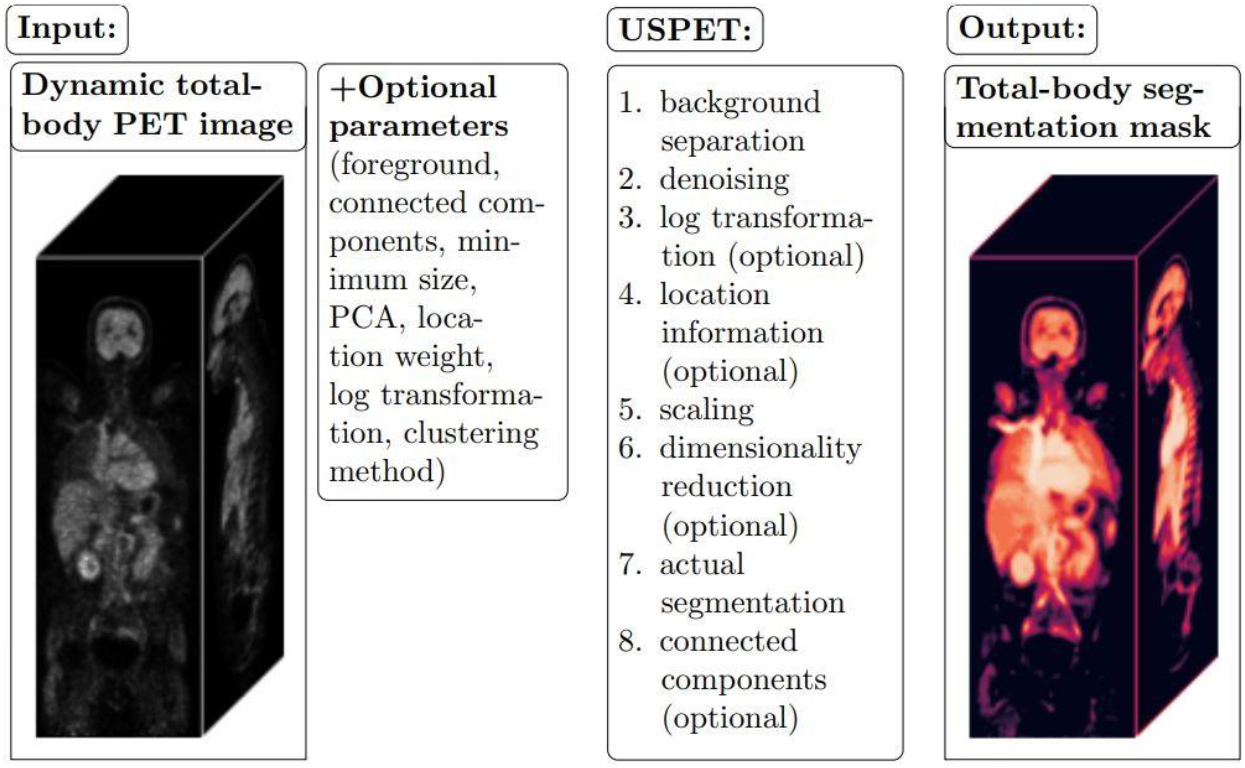
USPET pipeline. PCA refers to dimensionality reduction method principal component analysis.

Using a computer with 16GB of RAM and Intel Pentium Gold processor G6405T (CPU 3.50 GHz), analysing one image from the H_2_O dataset took on average approximately 40 minutes. The postprocessing step where tiny segments are merged into their surrounding bigger segments was the most time consuming part of the pipeline. The images in the FDG dataset had higher resolution (440· 440 · 354 vs 220 · 220 · 380 voxels within a similar scanned area) and it took approximately four hours to analyse one FDG image.

### 2.3. Test design

This study includes three different demonstrations of the strengths of USPET. The first one is a comparison of functional segments by USPET to manually drawn anatomical organs. We created the manual segments by initially automatically extracting them from the corresponding CT images using TotalSegmentator [18] (version 2.4.0), and then re-slicing them into the PET space. Typically, the participant had slightly moved during the long dynamic PET scan, so there was a mismatch between the PET and CT image. Those errors, together with the inaccuracies of the TotalSegmentator outputs, were then manually corrected in the Carimas software [19] (version 2.10) according to instructions from a medical expert with a degree in human anatomy. The match between USPET segments and manually drawn anatomical segments is measured using Jaccard index defined as the ratio of overlap ∩ and union ∪ of the segments:

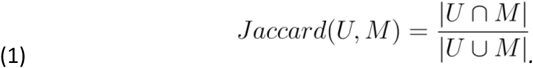

where |*U* ∩*M*| and |*U* ∪*M*| indicate the number of voxels in the overlap and union of the USPET segment (*U*) and the manual segment (*M*), respectively. Jaccard index is 1, if the two segments are identical, and 0 if they do not overlap at all. The higher the Jaccard index, the better the match.

Although segmentation is supposed to summarise the image, it is important that details relevant to the study are not lost in the process. Thus, in the second test we investigated whether the previously detected brain perfusion abnormalities can be seen from the segmentation results.

The strength of PET imaging is its ability to visualise functional processes instead of anatomically similar organs, and thus the segmentation results on PET alone are not expected to fully align with segments from anatomical imaging modalities such as CT or MRI. However, clinically meaningful segments should consider both aspects. Therefore, we investigated how the USPET segmentation works, if it is done within one anatomical organ only. This was carried out using the manually drawn organ masks from the FDG data for the heart, kidneys (left and right combined), and brain as the foreground for USPET. Then, the segmentation was done using three clusters for each organ. The number of clusters is not fine-tuned for the organs. Although a ground truth for these results does not exist, we demonstrate that the time activity curves of the PET image differ between the segments and that the output segments align with known functionally distinct parts of those organs.

## 3. Results

### 3.1. USPET vs anatomical segments

We compared the USPET segments to manually drawn anatomical segments for the brain, aorta, left and right kidney, liver, lungs, and heart using the Jaccard index as a measure of agreement. The mean and standard deviations of the Jaccard indices per organ are listed in Table 1. Unsurprisingly, homogeneous organs such as the brain, liver, and lungs had high mean Jaccards of 0.82, 0.72, and 0.70, respectively. As the kidneys are similar to each other and include functionally distinct areas, namely the medulla and cortex, the Jaccards between functional USPET segments and anatomical manual segments were low. Combining the right and left kidneys into one segment increased the mean Jaccard to 0.49. As the aorta and heart include high blood volume, they typically segment together in USPET, lowering the Jaccard indices of these organs, particularly for the aorta, as it is a smaller segment than the heart.

**Table 1.** Mean Jaccard indices and their standard deviations for each organ rounded to two decimals. The Jaccards are calculated between manually drawn anatomical segments and the best matching USPET segment from functional PET images.

| Organ | Mean | Standard deviation |
| --- | --- | --- |
| Brain | 0.82 | 0.02 |
| Aorta | 0.16 | 0.05 |
| Left kidney | 0.40 | 0.08 |
| Right kidney | 0.25 | 0.08 |
| Liver | 0.72 | 0.05 |
| Lungs | 0.70 | 0.05 |
| Heart | 0.57 | 0.08 |

### 3.2. Retaining of important details

While the level of detail in the segments is heavily dependent on their number and the used denoising, small yet relevant alterations in the original PET images can also be seen in the USPET segments even with reasonably few (here 100) segments considering the heterogeneity of a total-body image. The H_2_O dataset includes nine patients with abnormalities in brain PET images identified by manually inspecting the brain area of the images. Figure 2 illustrates that the total-body segmentation preserves these important changes even when they are subtle (e.g., subfigure 2A).

**Figure 2.**
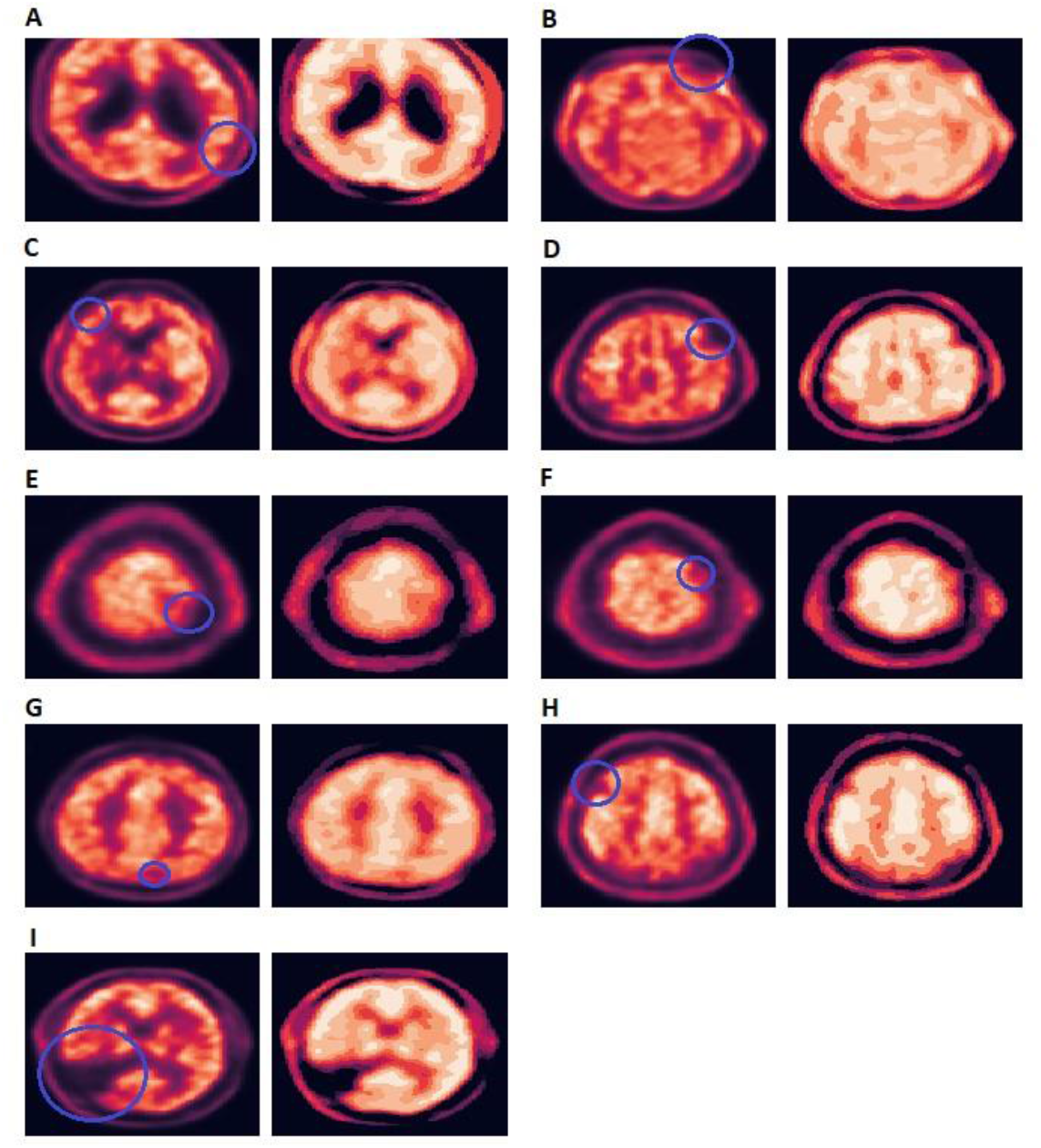
Axial slice of the original H_2_O PET scan with the suspected perfusion defect indicated with a blue circle (left-hand side) and the corresponding slice of the USPET segmented image (right-hand side) in the nine suspected cases (A-I). Notably, in this 2D visualisation there are some seemingly unexpected low activity areas not highlighted with the circles, but those are due to asymmetric positioning of the head during the scan or age-related brain atrophy, and not related to any health condition to our knowledge.

### 3.3. Combining anatomical and functional segmentation

While utilising CT-based anatomical segments is not yet an integrated part of the USPET pipeline, we wanted to test combining these two image modalities due to its relevance for clinical use. Here we manually investigate the PET-based segments by USPET, if the segmentation is done only within one anatomical organ using 3 segments. We used the brain, kidneys (both combined into one mask), and heart from the FDG dataset as test organs, because they include functionally distinct areas that are difficult to separate from a CT image.

The USPET heart segments are somewhat in agreement with the blood pool, the myocardium of the left ventricle, and the myocardium of the right ventricle. The kidneys are segmented into renal pelvis, cortex plus medulla, and a thin spillover area around the organs. USPET typically segments the brain roughly into white matter (plus ventricles and spinal fluid), grey matter, and spillover areas between these two, including also some lower activity brain regions. The time activity curves of the three segments have distinct profiles in each organ. Figure 3 illustrates the time activity curves of the three within-organ segments, and a randomly selected example of the segments obtained.

**Figure 3.**
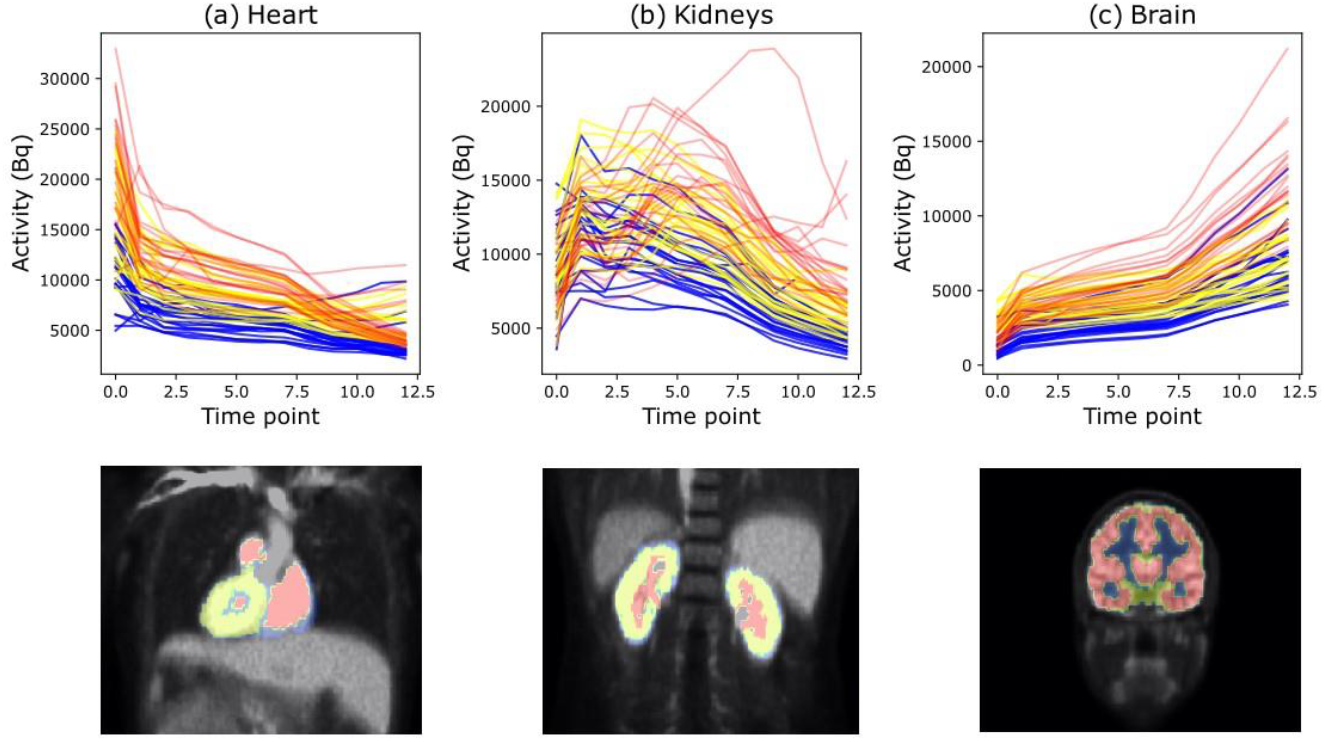
Three within-organ USPET segments labelled with red, yellow, or blue colour. The time activity curves are extracted from all analysed FDG images and one randomly selected segment example is visualised underneath.

## 4. Discussion and conclusions

In this paper, our aim was to develop a robust unsupervised segmentation method that works on modern PET images with a large field-ofview, and provide its implementation. We introduced USPET, which provides functionally meaningful segments that can be combined with anatomical segments obtained from the corresponding CT or MRI images for a good overview of the scanned body. Although the segments summarise the image and are therefore more coarse level information than the original PET image, USPET still preserves clinically important details, as demonstrated with suspected perfusion defects in the H_2_O dataset.

USPET is designed for modern dynamic total-body PET images, and thus its primary advantage is its capability to handle very large images without requiring heavy computational resources. Despite this lightness of the algorithm, the relevance of the segmentation results is not compromised, as demonstrated in this study. Its other strengths include a publicly available ready-to-use Python implementation, applicability to diverse images (e.g. different radiotracers, scanner sizes, organisms, time frame setups, etc.) and novelty. At the time of writing, we did not find a usable implementation of any other unsupervised general-purpose segmentation tool for large, modern PET image sets.

Besides the advantages listed above, USPET also has weaknesses. The most important is the slowness of one postprocessing step. The connected component analysis of ready segments generates thousands of small components, most of which need to be melted into their background segments within the USPET pipeline. However, this cleaning process is currently very time consuming, and, for example, in this study, we did not use the connected component post-processing with FDG dataset for time reasons. Also, this study itself has one main limitation: the difficulty of objective validation. Due to the lack of existing ground truth, the validation is done by demonstrating the biological and/or clinical relevance of the segments obtained.

Future work to further develop USPET pipeline include 1) code improvements to make the post-processing of connected component segments faster, 2) integrated option to utilise also CT image in the segmentation process, and 3) improving the Carimas plugin and similar implementations to other visualisation softwares favoured by clinicians. If the CT segments in improvement 2 are obtained from a supervised method (such as MOOSE [20] or TotalSegmentator [18]), the biological interpretation of the results becomes also simpler, as completely unsupervised approaches provide unlabelled segments.

## Data Availability

USPET is available as a Python implementation from github (https://github.com/rklen/USPET). The patient data is not available due to privacy restrictions.

## 5. Declarations

## 5.1. Acknowledgements

The authors thank David Ekwe for lending us his anatomy knowledge to do the manual segmentation of the FDG dataset.

## 5.4. Author contributions

The author contributions according to the CRediT classification are MKJ: conceptualisation, investigation, methodology, software, visualisation, writing (original draft preparation), HK: resources, data curation, TS: resources, OR: visualisation, AL: writing (original draft preparation), JK: resources, KAV: resources, RK: conceptualisation, supervision. All authors: writing - review and editing.

## 5.6. Informed consent and ethical approval

All patients were at least 18 years old, gave their written consent to the research use of their data, and the research from their data was approved by the Ethics Committee of the wellbeing services county of Southwest Finland.

## References

[1] Inês Domingues, Gisèle Pereira, Pedro Martins, Hugo Duarte, João Santos, and Pedro Henriques Abreu. Using deep learning techniques in medical imaging: a systematic review of applications on ct and pet. Artificial Intelligence Review, 53:4093–4160, 2020.

[2] Ivan S Klyuzhin, Guillaume Chaussé, Ingrid Bloise, Sara Harsini, Juan Lavista Ferres, Carlos Uribe, and Arman Rahmim. Psma-hornet: Fully-automated, multi-target segmentation of healthy organs in psma pet/ct images. Medical Physics, 51(2):1203–1216, 2024.

[3] Saeid Asgari Taghanaki, Yefeng Zheng, S Kevin Zhou, Bogdan Georgescu, Puneet Sharma, Daguang Xu, Dorin Comaniciu, and Ghassan Hamarneh. Combo loss: Handling input and output imbalance in multi-organ segmentation. Computerized Medical Imaging and Graphics, 75:24–33, 2019.

[4] Julian Leube, Matthias Horn, Philipp E Hartrampf, Andreas K Buck, Michael Lassmann, and Johannes Tran-Gia. Psma-pet improves deep learningbased automated ct kidney segmentation. Zeitschrift Für Medizinische Physik, 34(2):231–241, 2024.

[5] Silin Ren, Priscille Laub, Yihuan Lu, Mika Naganawa, and Richard E Carson. Atlas-based multiorgan segmentation for dynamic abdominal pet. IEEE Transactions on Radiation and Plasma Medical Sciences, 4(1):50–62, 2019.

[6] Albert Comelli. Fully 3d active surface with machine learning for pet image segmentation. Journal of Imaging, 6(11):113, 2020.

[7] Mhd Saeed Sharif, Maysam Abbod, Abbes Amira, and Habib Zaidi. Artificial neural network-based system for pet volume segmentation. Journal of Biomedical Imaging, 2010:1–11, 2010.

[8] Fereshteh Yousefirizi, Isaac Shiri, Joo Hyun O, Ingrid Bloise, Patrick Martineau, Don Wilson, François Bénard, Laurie H Sehn, Kerry J Savage, Habib Zaidi, et al. Semi-supervised learning towards automated segmentation of pet images with limited annotations: application to lymphoma patients. Physical and Engineering Sciences in Medicine, 47(3):833–849, 2024.

[9] Runxi Cui, Zhigang Chen, Jia Wu, YanLin Tan, and GengHua Yu. A multiprocessing scheme for pet image pre-screening, noise reduction, segmentation and lesion partitioning. IEEE Journal of Biomedical and Health Informatics, 25(5):1699–1711, 2020.

[10] Jacob W Vogel, Niklas Mattsson, Yasser Iturria-Medina, Olof T Strandberg, Michael Scöll, Christian Dansereau, Sylvia Villeneuve, Wiesje M van der Flier, Philip Scheltens, Pierre Bellec, et al. Data-driven approaches for tau-pet imaging biomarkers in alzheimer’s disease. Human brain mapping, 40(2):638– 651, 2019.

[11] Ziyue Xu, Mingchen Gao, Georgios Z Papadakis, Brian Luna, Sanjay Jain, Daniel J Mollura, and Ulas Bagci. Joint solution for pet image segmentation, denoising, and partial volume correction. Medical image analysis, 46:229–243, 2018.

[12] Jinman Kim, Weidong Cai, Dagan Feng, and Stefan Eberl. Segmentation of voi from multidimensional dynamic pet images by integrating spatial and temporal features. IEEE Transactions on Information Technology in Biomedicine, 10(4):637–646, 2006.

[13] Sandrine Mouysset, Hiba Zbib, Simon Stute, Jean-Marc Girault, Jamal Charara, Joseph Noailles, Sylvie Chalon, Irène Buvat, and Clovis Tauber. Segmentation of dynamic pet images with kinetic spectral clustering. Physics in Medicine & Biology, 58(19):6931, 2013.

[14] Brian J Parker and Dagan Feng. Graph-based mumford-shah segmentation of dynamic pet with application to input function estimation. IEEE Transactions on Nuclear Science, 52(1):79–89, 2005.

[15] Renaud Maroy, Raphäel Boisgard, Claude Comtat, Vincent Frouin, Pascal Cathier, Edouard Duchesnay, Frédéric Dollé, Peter E Nielsen, Régine Trébossen, and Bertrand Tavitian. Segmentation of rodent whole-body dynamic pet images: an unsupervised method based on voxel dynamics. IEEE transactions on medical imaging, 27(3):342–354, 2008.

[16] Jinxiu Cheng-Liao and Jinyi Qi. Segmentation of mouse dynamic pet images using a multiphase level set method. Physics in Medicine & Biology, 55(21):6549, 2010.

[17] Maria K Jaakkola, Maria Rantala, Anna Jalo, Teemu Saari, Jaakko Hentilä, Jatta S Helin, Tuuli A Nissinen, Olli Eskola, Johan Rajander, Kirsi A Virtanen, Jarna C Hannukainen, Francisco López-Picón, and Riku Klén. Segmentation of dynamic total-body [18f]-fdg pet images using unsupervised clustering. International Journal of Biomedical Imaging, 2023:3819587, 2023.

[18] Jakob Wasserthal, Hanns-Christian Breit, Manfred T Meyer, Maurice Pradella, Daniel Hinck, Alexander W Sauter, Tobias Heye, Daniel T Boll, Joshy Cyriac, Shan Yang, et al. Totalsegmentator: Robust segmentation of 104 anatomic structures in ct images. Radiology: Artificial Intelligence, 5(5), 2023.

[19] Oona Rainio, Chunlei Han, Jarmo Teuho, Sergey V Nesterov, Vesa Oikonen, Sauli Piirola, Timo Laitinen, Marko Tättäläinen, Juhani Knuuti, and Riku Klén. Carimas: an extensive medical imaging data processing tool for research. Journal of Digital Imaging, pages 1–9, 2023.

[20] Lalith Kumar Shiyam Sundar, Josef Yu, Otto Muzik, Oana C Kulterer, Barbara Fueger, Daria Kifjak, Thomas Nakuz, Hyung Min Shin, Annika Katharina Sima, Daniela Kitzmantl, et al. Fully automated, semantic segmentation of whole-body 18f-fdg pet/ct images based on data-centric artificial intelligence. Journal of Nuclear Medicine, 63(12):1941–1948, 2022.

